# Acupuncture for the treatment of pregnancy-related low back pain: a systematic review and network meta-analysis

**DOI:** 10.1101/2023.11.09.23298330

**Authors:** Min Li, Zongyi Xiao, Dongling Tan, Daqiang Zhao, Qi Chen

**Author notes:** These authors contributed equally to this work (Min Li, Zongyi Xiao, Dongling Tan). Correspondence: Daqiang Zhao Department of Anesthesiology, Sichuan Taikang Hospital, Chengdu, People’s Republic of China, Correspondence: Qi Chen Department of Anesthesiology, Chongqing University Cancer Hospital, Chongqing, People’s Republic of China.

## Abstract

**Background:** Despite the effectiveness of acupuncture in the treatment of musculoskeletal pain, many physical therapists are unwilling to use it on pregnant women. A recent systematic review of acupuncture for pregnant women did not include a comparison with sham acupuncture (SAcu). Thus, we aimed to explore the effects of acupuncture, SAcu, and standard care (SC) on pregnancy-related low back pain.

**Methods:** We searched five different medical literature databases (PubMed, Embase, MEDLINE, Springer, and Google Scholar) from inception to September 30, 2022. After screening, the following methods were identified: acupuncture, SAcu, and SC. The primary outcome was visual analog scale (VAS) intensity after the intervention. The secondary outcomes were the overall effects of treatment, quality of life (QOL), and QOL evaluated using the Short Form-36 Health Survey Questionnaire (SF-36).

**Results:** The network meta-analysis included eight studies and 864 patients. Acupuncture and SAcu were relatively more advantageous in terms of analgesic effects after intervention than SC, but there were no differences between them. In terms of overall effects in number of remissions and the SF-36, Acupuncture was found to be superior to other methods, and SAcu was better than SC. Acupuncture had the highest surface under the cumulative ranking curve, followed by SAcu and SC for all outcomes.

**Conclusions:** Acupuncture performs similarly to SAcu in pain relief and is more efficient than SC. Regarding the effectiveness of treatment and QOL, acupuncture therapy was superior to SAcu and SC.

## Background

Low back pain (LBP) and pelvic girdle pain (PGP) during pregnancy are common and characterized by musculoskeletal pain located between the hip crease and the ribs, with or without leg pain, with a prevalence ranging from 45–77%.^1–4^ LBP usually starts at 12–24 weeks and reaches its maximum intensity from weeks 24–36 in most pregnant women.^1^ Symptoms are usually milder in the morning and worsen at night. Strenuous work, standing, and physical exercise involving the lower back and pelvis can aggravate LBP, which affects work or other daily activities to a large extent.^5,6^

Some interventions that have been proposed include education, pelvic belts, physiotherapy, exercises, pharmacological therapy, transcutaneous nerve stimulation, and acupuncture,^7^ but the effects remain to be discussed. In 2015, the American Congress of Obstetricians and Gynecologists (ACOG)^8^ suggested that moderate-intensity (Borg 13-14) exercise should be encouraged for all women with uncomplicated pregnancies during pregnancy, which could strengthen the abdominal and back muscles and minimize the risk on the low back. Nevertheless, adherence to exercise is difficult with advancing pregnancy; inappropriate exercise posture may lead to instability and misalignment, resulting in an increased load on the affected joints and ligaments, causing micro-damage or unacceptable falls, and threatening the fetus. Acupuncture originated from traditional Chinese medicine and has been used for the prevention and treatment of pain for millennia. Since it was introduced to the West centuries ago, new styles and forms of acupuncture have developed, including hand, foot, scalp, ear (auricular), and body acupuncture. The most common method is to use a thin, solid, metallic needle to gently penetrate and rotate into the trigger points at the site or in the vicinity of pain. Accordingly, the patients get a needle sensation or Dechi, often expressed as numbness, dullness, or tingling.

As reported in some studies,^2,9,10–19^ acupuncture therapy has a significant benefit in pain relief in most participants. Despite the effectiveness of acupuncture in the treatment of musculoskeletal pain, many physical therapists are unwilling to use it in pregnant women, as they believe that some points might trigger uterine contractions, inducing preterm labor, associated with the risk of litigation.^2^ A recent systematic review^20^ involving 10 studies with 1040 women with LBP and/or PGP during pregnancy showed that compared with standard care (SC), acupuncture was associated with pain relief, functional status, and quality of life (QOL) improvement. Moreover, no severe adverse effects were observed in the newborns. However, it did not include a comparison with sham acupuncture (SAcu; inserting needles at non-acupuncture points). As we know, pain is subjective; SAcu may ensure participants are blind, the potential bias that could affect the outcomes were reduced, and the possible mechanisms of acupuncture effects could be better known. It is necessary to reassess the existing evidence, which prompted us to conduct a network meta-analysis to compare the effectiveness of acupuncture, SAcu, and SC on pain relief and treatment of LBP.

## Methods

### Data sources and searches

Following the Preferred Reporting Items for Systematic Reviews and Meta-Analyses (PRISMA) guidelines, this network meta-analysis with systematic review was registered at the International Prospective Register of Systematic Reviews (PROSPERO) with registration number CRD42022364064. English-language studies were searched mainly through five databases (PubMed, Embase, Medline, Springer, and Google Scholar) from inception to September 30, 2022. Various combinations, keywords, and MeSH terms were used, using ‘low-back pain,’ ‘pelvic pain,’ ‘pelvic girdle pain,’ ‘acupuncture,’ ‘sham acupuncture,’ ‘standard care,’ ‘ear acupuncture,’ ‘pregnancy,’ ‘pregnancy-related,’ ‘lumbar back pain,’ ‘posterior pelvic pain,’ ‘standard treatment,’ ‘auricular acupuncture,’ or ‘anesthesia, general.’

### Study selection and quality assessment

We included randomized controlled trials (RCTs) of pregnant women with LBP with or without PGP who were treated with acupuncture, SAcu, or SC, regardless of different acupuncture points or needle materials. Cohort studies, case reports, animal studies, letters, and other complications during pregnancy such as eclampsia, inflammation, impaired nerve function, or LBP before pregnancy were excluded. All studies were imported into Endnote X9 (Clarivate, London, UK) by two researchers (ML and ZYX), and duplicate studies were removed. For the references that seemed to meet the inclusion criteria, the full text was reviewed to determine the final selection.

The Cochrane risk of bias (ROB) assessment tool was used to assess article quality by two researchers (ML and ZYX), which included random sequence generation, allocation concealment, blinding, integrity of outcome data, selective reporting of results, and other biases. Each aspect was classified into three levels (low, unclear, or high risk). If there was any dispute, a third investigator (DQC) provided consultation until a consensus was reached.

### Outcome measures and data extraction

Two investigators (ML and ZYX) independently extracted the data. If data were reported as median (interquartile range), software (https://www.math.hkbu.edu.hk/~tongt/ papers/median2mean.html) was used to obtain the mean and standard deviation. If the data were represented only in a graphical format, they were numerically extrapolated by plot digitization using Plot Digitizer 2.6.8, (Free Software Foundation). The extracted data included authors, publication time, sample size, gestational age, intervention point, and outcome measures (Table 1). The primary outcome was the pain intensity after the intervention. The visual analog scale (VAS) was used to evaluate changes in pain intensity (using a scale from 0 to 10 cm, where 0 is no pain and 10 is the worst pain). The secondary outcomes were the overall effects of treatment and QOL (participants with a VAS score lower than 30% after intervention). The QOL was expressed by the Short Form-36 Health Survey Questionnaire (SF-36).

**Table 1.** Characteristics.

| Study ID | Year | Sample size<br>Acu/SAcu//SC | Gestation<br>age | Intervention point | Outcome |
| --- | --- | --- | --- | --- | --- |
| Foster et.al | 2016 | 42/42//41 | more than<br>24 weeks | BL25, BL26,<br>BL31, BL54 | 1, 2, 3 |
| Vas et. al | 2019 | 55/55//52 | 24-36<br>weeks | Shenmen and<br>Kidney points | 1, 2, 3 |
| Wedenberg<br>et. al | 2000 | 28/0//18 | no more<br>than 32<br>weeks | Bl:26–30, Bl: 60,<br>CW 2 | 1, 2 |
| Silva et. al | 2004 | 27/0//34 | 15 to 30<br>weeks, | K13, S13, BL62,<br>BL40,TE5,<br>GB30,GB41,<br>huatojiaji points | 2 |
| Nicolianl et al | 2019 | 96/0//103 | 16 and 34 weeks | Weizhong, AShi points | 1, 2 |
| Kvorning et. al | 2004 | 37/0//35 | 24-37 weeks | BL60, SI3, BL22–26, GV20 | 2 |
| Wang et. al | 2009 | 58/54//47 | 25 to 38 weeks | CW8, SC7, TF2 points | 1, 2, 3 |
| Ekdahl et. al | 2010 | 20/20//0 | 20-26 weeks | Known anatomical site as reference points | 1, 3 |

Acu: Acupuncture, SC: Standard care. 1: Pain scores after treatment; 2: Overall effects of treatment, 3:
Quality of life (Short Form-36 Health Survey Questionnaire (SF-36))

### Statistical analyses

The data were analyzed using R version 4.0 (R Foundation, Vienna, Austria). The consistency of the model was also assessed, with *P* > 0.05 indicating good consistency between the direct and indirect results. Odds ratios and 95% confidence intervals (CIs) served as the effect indicators for dichotomous outcomes, whereas the mean difference and 95% CIs were used for continuous outcomes. A network plot and ranking diagram were drawn for each intervention. Surfaces under the cumulative ranking curves (SUCRAs) were used to rank intervention results. The SUCRA reflects the merits and defects of interventions. When it is close to one, the intervention is more effective. Finally, the results were shown using forest maps. Statistical significance was set at *P* < 0.05. Publication bias was examined using comparison-adjusted funnel plots.

## Results

### Study selection

A total of 646 articles were obtained through the preliminary screening. After removing 38 duplicate articles, 608 trials remained. After reading the title and abstract, 518 articles were excluded because there was no control or placebo group or because it belonged to a case report, meeting abstract, or review. Ninety full-text articles were assessed for eligibility. Finally, 8 articles were included in the qualitative and quantitative analyses. A flowchart is shown in Fig. 1. VAS after intervention was reported in six of these RCTs, while seven RCTs reported overall effects of treatment and four RCTs reported SF-36 results.

**Fig. 1:**
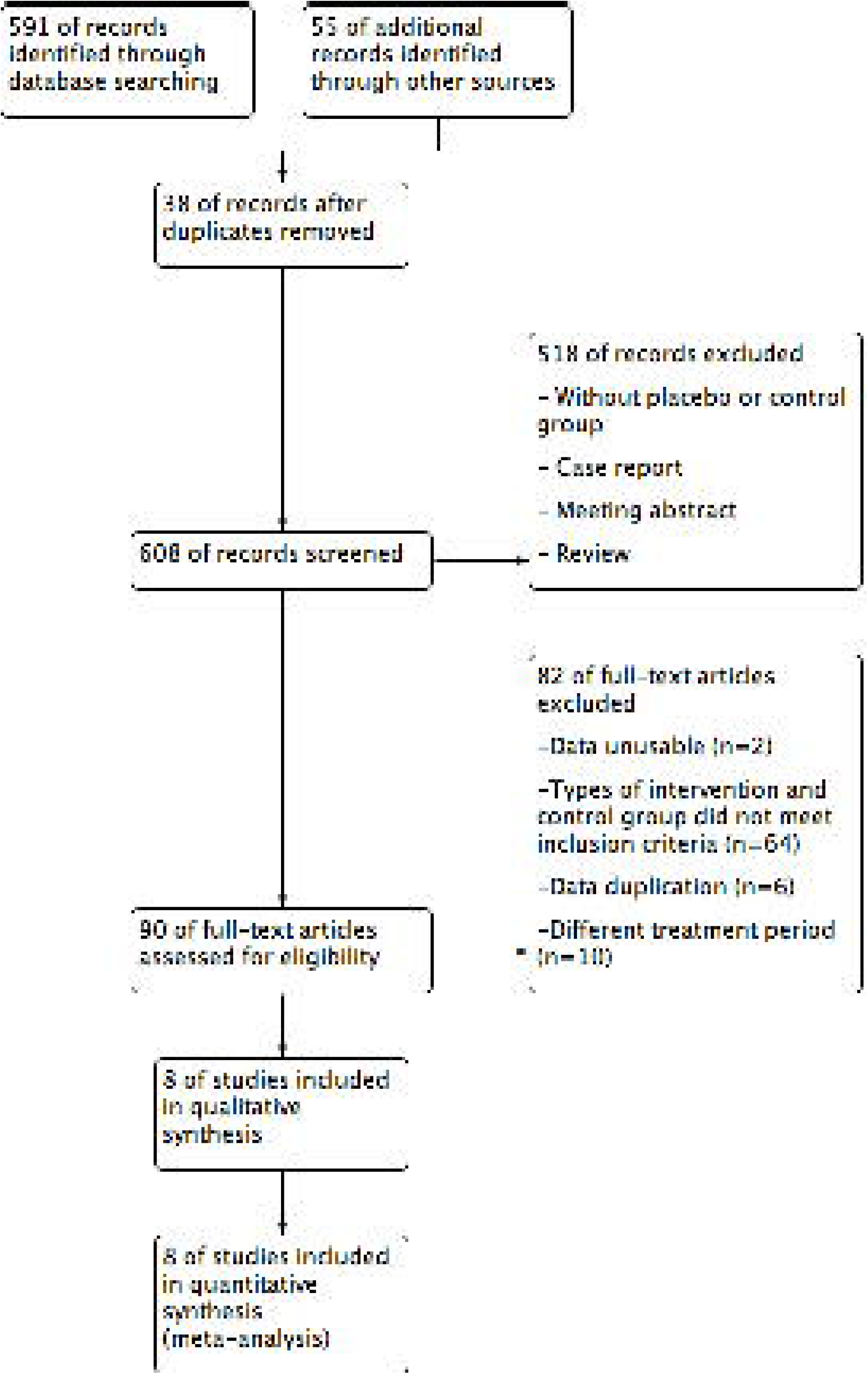
The screening flow chart.

Three studies compared the difference between acupuncture, SAcu, and SC directly,^15,17,18^ four studies only compared the difference between acupuncture and SC,^2,9,14,16^ and one study compared acupuncture and SAcu^19^ (Fig. 2). The basic features of the included studies are presented in Table 1.

**Fig. 2:**
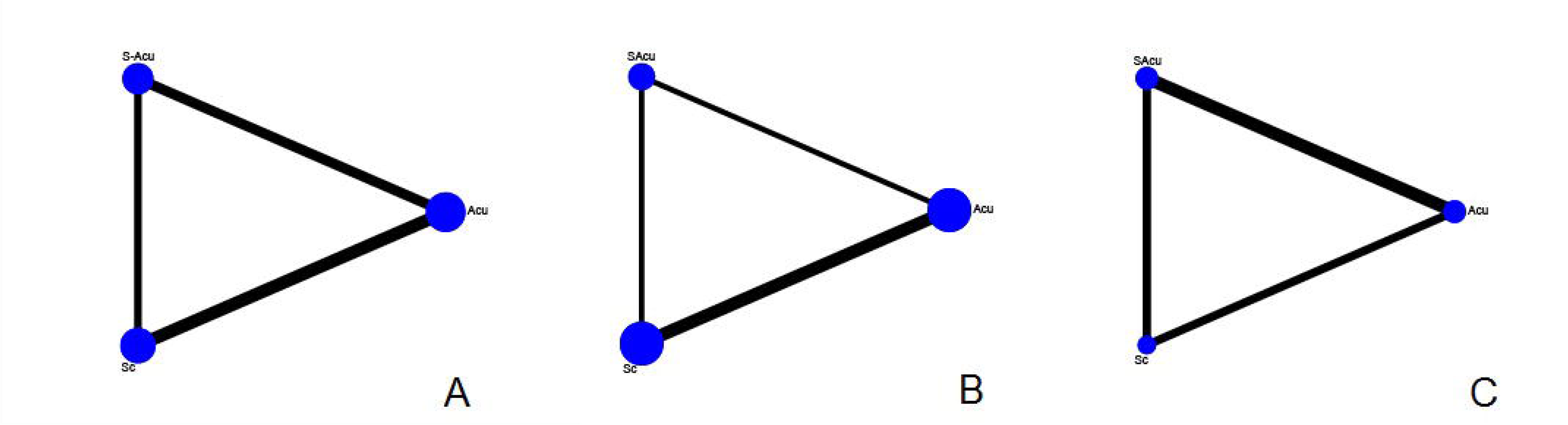
Evidence networks. Acu: Acupuncture, SAcu: Sham Acupuncture, SC: Standard care. A: Pain scores after treatment; B: Overall effects of treatment; C: Quality of life. The lines represent direct comparison of interventions, the line thickness represents the number of studies, and the dot size represents the sample size.

### Risk of bias assessment

Six studies had a low risk of bias^9,14–18^ and two studies had a high risk of bias.^2,19^ This bias mainly came from allocation concealment and blinding. Seven studies randomly assigned groups using computer software or drawing lots,^9,14–19^ and one study used the admission time to assign participants to groups.^2^ The allocation of six trials was concealed,^9,14,15,17,18^ while that of two trials was unclear.^2,19^ All outcome data were complete (Fig. 3).

**Fig. 3:**
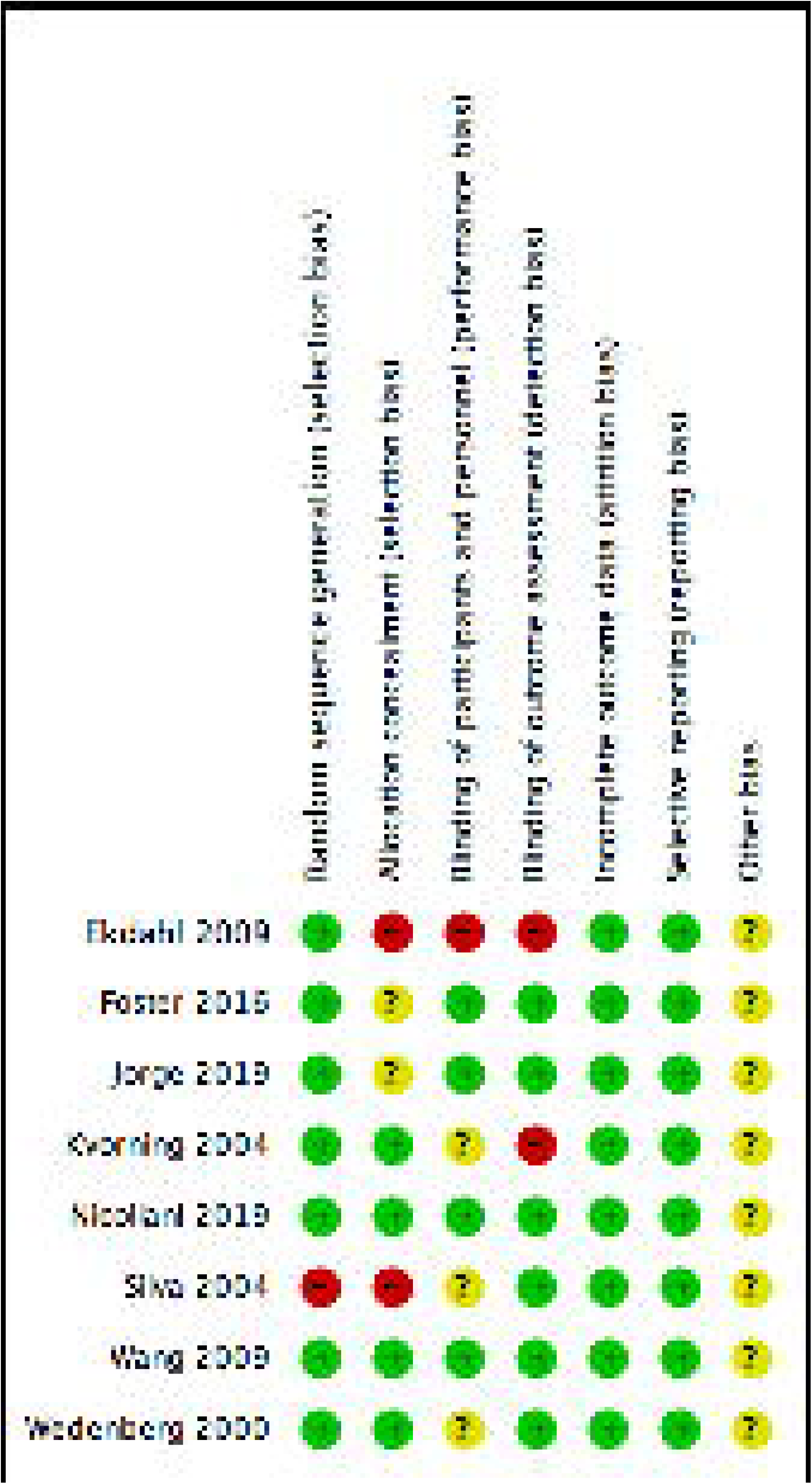
Risk of bias analysis. Green means low risk of bias, yellow means unclear, and red means high risk of bias.

### VAS scores after intervention

VAS scores after intervention were reported in 731 participants in six RCTs.^9,15–19^ Compared with the SC group, the VAS scores of the acupuncture and SAcu groups were significantly lower (*P* < 0.05). Meanwhile, the VAS score of the acupuncture group was lower than that of the SAcu group by 0.75, but the difference between them was not significant (*P* > 0.05) (Fig. 4A). The suggested rank order probability for better analgesia after the intervention was acupuncture > SAcu > SC (Fig. 5A).

**Fig. 4:**
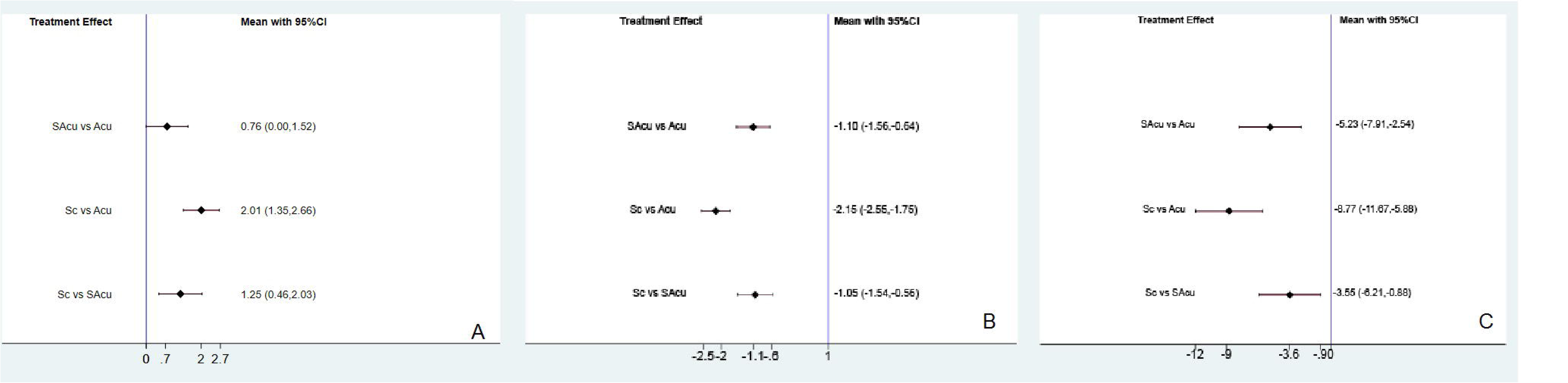
Forest maps. Acu: Acupuncture, SAcu: Sham Acupuncture, SC: Standard care. A: Pain scores after treatment; B: Overall effects of treatment; C: Quality of life.

**Fig. 5:**
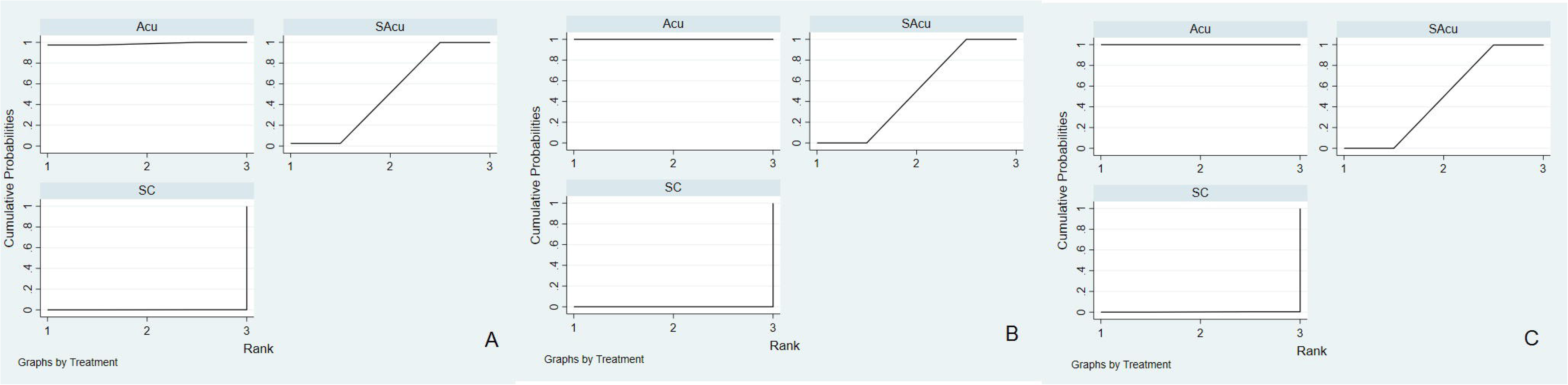
SUCRA for different interventions. Acu: Acupuncture, SAcu: Sham Acupuncture, SC: Standard care. A: Pain scores after treatment; B: Overall effects of treatment; C: Quality of life.

### Overall effects of treatment

The overall effects of treatment after intervention were reported in 824 participants in seven RCTs.^2,9,14,15–18^ The overall effects of the acupuncture and SAcu groups were greater than those of the SC group (*P* < 0.05). The effect of the acupuncture group was significantly higher than that of the SAcu group (*P* < 0.05) (Fig. 4B). The suggested rank order probability for better analgesia after the intervention was acupuncture > SAcu > SC (Fig. 5B).

### Quality of life

QOL was reported in 486 participants in four RCTs.^15,17–19^ The QOL of the acupuncture and SAcu groups improved after the intervention compared to the SC group (Fig. 4C). The QOL of the acupuncture group was significantly better than that of the SAcu group (*P* < 0.05). The suggested rank order probability for better analgesia after the intervention was acupuncture > SAcu > SC (Fig. 5C).

## Discussion

With respect to pain reduction, the overall results indicated that acupuncture therapy was similar to SAcu therapy and more efficient than SC; acupuncture therapy was superior to the other two groups in terms of the effectiveness of treatment and QOL.

LBP is a common syndrome during pregnancy, and the underlying mechanisms of its etiology remain unknown. On the one hand, due to the imbalance between the pelvis and the lumbar spine segments,^1^ excessive lordosis is caused by increased uterine volume.^2^ Relaxin (a peptide hormone found in the placenta, corpus luteum, and decidua of pregnant women) can relax the pubic symphysis and sacroiliac ligaments.^3^ The compression of the lumbosacral nerve roots by the fetus and the reduction of blood flow caused by the compression of large vessels by the pregnant uterus also plays an important role in the occurrence of LBP.^2^ LBP subsides shortly after delivery in most women.^21^ Nevertheless, approximately 43% of mothers continue to experience constant pain 6 months after delivery, including 7% with recurrent pain and 36% with persistent pain.^22^ Moreover, 20% of pregnant women with LBP reported persistent complaints three years after delivery.^23^

Exercise, pharmacological therapy, and acupuncture are the most common interventions used in clinical practice. Acetaminophen^24,25^ is considered safe and is the first choice of pharmacotherapy to relieve pain during pregnancy. However, because the maternal-fetal circulation is unique and the drugs may have potential effects on the fetus, drug therapy during pregnancy remains challenging.^26^ In previous meta-analyses,^7^ the effect of physiotherapeutic interventions on pregnancy-related lumbopelvic pain has been discussed, and there was strong evidence for the positive effects of acupuncture compared with exercise in general and for specific stabilizing exercises like water gymnastics. The underlying mechanism of action of acupuncture is not completely understood. Existing research suggests the following theories. First, peripheral and central pain control systems were activated by acupuncture through the release of different endogenous opioid or non-opioid compounds, such as β-endorphin, enkephalin, γ-aminobutyric acid, deilorphine, serotonin, norepinephrine or ATP, to exert analgesic effects.^9–12^ Secondly, the gate control theory of pain was considered to be how acupuncture works, in which the sensory input to the central nervous system was inhibited by the inserted needle. Third, the vascular and immunomodulatory factors, such as inflammatory mediators, was stimulated by the presence of the needle as a foreign substance within the body tissue to reduce pain.^13^ However, because the appropriate position and manipulation of the needles are considered essential in achieving successful outcomes, acupuncture applications are restricted and need specialized personnel.^7,27,28^

Yao et al ^29^ and Yang et al ^20^ showed that acupuncture was superior to the treatment provided to the control group. However, they focused on evaluating the efficacy of acupuncture and SC; SAcu was not involved and there was no appropriate blinding. The acupoints in SAcu may not be in the appropriate or specific place for treatment, and SAcu is generally designed to achieve good credibility and blinding and to minimize the potential physiological effects. In our review, both acupuncture and SAcu exerted non-specific effects on pain relief compared with SC, which is consistent with previous studies.^30^ To the best of our knowledge, there are several possible reasons for this phenomenon. First, the needle touching the skin can possibly be considered as a type of sensory stimulation, which could activate mechanoreceptors coupled to slow-conducting unmyelinated (C) nerve fibers and consequently expresses physiological responses. Therefore, any specific acupuncture intervention may cause physiological responses and pain relief.^31,32^ Second, patients’ expectations and therapeutic effects could be regulated by the underlying psychobiological mechanisms, which could trigger complex neurobiological changes in the central and peripheral nervous systems, as well as the end-organs, and elicit non-specific effects contributing to the overall therapeutic effect.^30,33–34^ In addition, pain relief may also be associated with long consultations during pregnancy that are in themselves prone to be psychologically therapeutic.^34–36^ Overall, results from SAcu suggest that the therapy is not completely inert. For this reason, acupuncture therapy may be applied by non-specialists and primary care professionals in a clinical context, thus bringing it closer and more convenient to patients. Meanwhile, it is not necessary for a patient to experience Dechi on each occasion, reduce the difficulty of acupuncture stimulation, and avoid unnecessary pain and complications. Experiencing pain involves a range of suffering for the individual, which may influence daily life. As we can see in the review, there are tendencies of improvements in the QOL for pregnant women with pain relief. Although small bruises and subcutaneous hematomas, transient ear tenderness that resolved spontaneously, and paresthesia in the arm have been reported in some trials,^2,15–17^ no serious consequences persisted. According to relevant studies,^20,37–38^ no significant side effects were observed in either the mother or the fetus, including obstetrical and neonatal morbidity, mean birthweight, and caesarean delivery rate.

This study had several limitations. First, there was heterogeneity in our review. We supposed that the methodological bias and differences could have resulted in this heterogeneity: two ear acupuncture RCTs and five acupuncture RCTs were included in the review, and the points of puncture, depth of puncture, duration of puncture, and gestational age at which the treatment was performed differed. Third, the measurement tools may not be the best choice; the VAS or the numeric rating scale play an important role in assessing the clinical pain intensity, but pain disorders are usually complex, multi-factorial, and have an incompletely understood pathophysiology. In future studies, using multi-dimensional pain assessment tools to address different aspects of pain could be taken into consideration, such as the Brief Pain Inventory^39^ or McGill Pain Scale.^40^ Considering the above limitations, evaluating the efficacy of acupuncture for LBP requires larger-scale, higher-quality evidence, and rigorously designed RCTs.

## Conclusion

Current evidence suggests that acupuncture therapy performs similarly to SAcu therapy in pain relief and is more efficient than SC. Acupuncture therapy is superior to SAcu and SC in terms of effectiveness of treatment and QOL.

### List of abbreviations

CI: confidence interval
LBP: low back pain
PGP: pelvic girdle pain
QOL: quality of life
RCT: randomized controlled trial
SAcu: sham acupuncture
SC: standard care
SF-36: Short Form-36 Health Survey Questionnaire
SUCRA: surface under the cumulative ranking curve
VAS: visual analog scale

## Declarations

## Ethics approval and consent to participate

Not applicable.

## Consent for publication

Not applicable.

## Availability of data and materials

All data are included in this article

## Authors’ contributions

Study concept and design: D.Z, D.T. Acquisition of data: M.L, Z.X, D.Z. Manuscript drafting: M.L, Z.X, D.T. methodology and software: Q.C, D.Z. Study supervision: D.Z, Q.C. All authors read and approved the final manuscript.

## Data Availability

All data produced in the present work are contained in the manuscript

## Acknowledgments

Not applicable.

